# What healthcare programs have been developed for the management of musculoskeletal pain in Indigenous populations? A scoping review

**DOI:** 10.1101/2025.04.06.25325313

**Authors:** Jane Linton, Ivan Lin, Veronica Matthews, Emma Walke, Eduardo da Silva Alves, Simon RE Davidson, Amanda Tutty, Christopher M Williams

## Abstract

**Objective:** There is a current knowledge gap and need for a comprehensive summary of literature about the management of musculoskeletal pain for Indigenous people internationally. We aimed to summarise and appraise cultural and methodological quality of studies describing programs developed to support Indigenous peoples manage musculoskeletal pain.

**Design:** Scoping review.

**Data sources:** Systematic literature search of Medline, Embase, CINAHL, Proquest Central and Scopus from January 1993 to November 2023.

**Eligibility criteria:** Scientific literature published in English; conducted in Canada, United States of America, Australia and New Zealand; describing the development and/or implementation of programs or components of programs for Indigenous patients with musculoskeletal pain or health staff working with Indigenous patients with pain musculoskeletal pain were included.

**Results:** 23 articles met the inclusion criteria. Articles were classified into two categories: programs for patients (n=16) and programs for health staff working with Indigenous peoples (n=7). Overall methodological quality of studies was moderate-high, overall cultural quality of studies was moderate. Most programs had positive outcomes and were well received by participants. Multiple strategies, including culturally appropriate models of care and locations, were incorporated into the programs and most studies considered cultural components for the development and delivery of the programs.

**Conclusion:** Although there has been a recent increase in research for the management of musculoskeletal pain amongst Indigenous peoples it is still limited. Cultural considerations are critical for research methodology and the development and delivery of new health programs focussed on improving the health and wellbeing of Indigenous peoples with musculoskeletal pain.

**What is already known in this topic:**

- Indigenous peoples internationally experience an inequitable burden due to musculoskeletal pain and have reduced access to culturally safe and appropriate health programs for musculoskeletal pain.
- Culturally safe and appropriate healthcare programs have demonstrated positive outcomes for Indigenous peoples with other chronic disease conditions.
- Little is known about the western methodological or cultural rigour of research, features, or outcomes of musculoskeletal programs for Indigenous peoples.

**What are the new findings:**

- Programs have been developed to improve the management of musculoskeletal pain for Indigenous peoples, by providing interventions for Indigenous peoples and education training for health staff working with Indigenous peoples with musculoskeletal pain conditions. A comprehensive summary of the programs can inform Indigenous Community members, clinicians and researchers in the future development of culturally safe, appropriate and accessible programs for Indigenous peoples.
- Strategies for the development and delivery of health programs, such as the co-design of programs with Indigenous peoples and the implementation of programs in culturally safe locations, have resulted in positive outcomes for Indigenous peoples with musculoskeletal pain conditions.
- Musculoskeletal health research with Indigenous peoples needs to be of a high quality from both methodological and cultural perspective. Examples of how to conduct or appraise future studies are demonstrated.

## INTRODUCTION

Internationally there is an increasing focus on reducing the burden of musculoskeletal conditions amongst Indigenous peoples and need to improve the equity in accessing healthcare programs.[1] Musculoskeletal conditions, described as a “group of conditions, along with arthritis and other conditions, that affects the bones, muscles and joints”,[2] are major contributors to health burden and disability.[1] Conditions, such as osteoarthritis and chronic pain, disproportionally affect Indigenous peoples worldwide [3, 4] and there are inadequate services available to provide appropriate care.[5]

The Indigenous peoples of Canada, Australia, Aotearoa (New Zealand) and United States of America (USA) (collectively known as CANZUS) have cared and looked after their people, Communities (see supplemental table 1 for terms used from an Indigenous context), land, waterways and animals for thousands of years through Indigenous methods and with culturally embedded practices. These practices align with Indigenous views of health and wellbeing, and the interconnectedness of physical, social, spiritual, cultural and Community wellbeing, including the health of environment and homelands. Health programs like Birthing on Country[6] and ‘Healthy Country, healthy people’[7] that embrace Indigenous holistic viewpoints have reported positive outcomes.

To date musculoskeletal research has focussed primarily in mainstream health care systems, and includes health programs that aim to improve function, mobility, reduce or learn to cope with pain.[8, 9] Recommended health programs for people with musculoskeletal pain include education and advice for self-management, physical activity, exercise, and the gradual and guided return to participation in leisure activities and sports.[10] Other research suggest targeting skills of health professionals who work with people with musculoskeletal pain, such as communication and health coaching is also effective.[11] Despite a substantial musculoskeletal pain research base in mainstream health care, few musculoskeletal programs for Indigenous peoples are reported that are developed and built upon Indigenous views of wellbeing. As a result, many Indigenous peoples faced with these conditions are unable to access care that respects their cultural and broader health needs.

Greater focus on cultural appropriate components of musculoskeletal pain services has the potential to overcome common barriers and improve accessibility for Indigenous peoples. Typical barriers include inflexible appointment attendance policies, culturally unsafe environments and communication with health staff.[3, 12] Addressing these through culturally informed communication and settings, working with trusted health staff, the inclusion of culturally appropriate resources and traditional healing practices have the potential to improve the experiences of care and health outcomes of Indigenous peoples by building on cultural strengths.[3, 13, 14]

Although there are robust guidelines about conducting and reporting musculoskeletal health research,[15, 16] there has been an increased focus internationally on how research is conducted with Indigenous Communities and what areas of health are being researched. Historically there has been a deficit discourse when Indigenous people have been the subject of health research undertaken by non-Indigenous researchers[14] and Indigenous Communities have had little involvement in the research process or outcomes.[17] Research conducted in this manner has resulted in unethical, poorly conducted and reported research that reinforces the negative narrative of Indigenous health inequalities and poorer health outcomes.[18] This method also excludes an Indigenous lens to interpret research data and prevents self-determination of Indigenous peoples to develop sustainable, Community driven solutions.[19] Recently, Indigenous health researchers have led the way to improve the appraisal and reporting standards of research practices from a cultural perspective when working with Indigenous peoples.[20–22] The Aboriginal and Torres Strait Islander Quality Assessment Tool (QAT) was developed to guide Australian reviews from an Aboriginal and Torres Strait Islander perspective.[21] Internationally, the CONSIDER statement has been endorsed as a guide for researchers conducting Indigenous health research.[20] Thus far these instruments have not been used to appraise musculoskeletal pain research with Indigenous peoples.

To date, most musculoskeletal pain research has been developed without the consideration of minority populations. With the emergence of research focused on Indigenous health by Indigenous researchers and Community members, there is broader recognition of best practice to conduct research and the benefits. Despite increased acknowledgement of the need for Indigenous research into musculoskeletal pain conditions and increased research activity [23, 24] there has been very little synthesis of the existing research to inform what is the current status and future directions. We aimed to summarise what culturally appropriate health programs exist for Indigenous peoples in the CANZUS region with musculoskeletal pain and appraise cultural and methodological considerations of the included studies.

## METHODS

This review was conducted in accordance with the updated JBI methodological guidance for the conduct of scoping reviews[25] and reported with the Preferred Reporting Items for Systematic Reviews and Meta-analyses Extension for Scoping Review (PRISMA-Scr) guide.[26] The protocol was registered on Open Science Framework.[27]

### Equity, diversity, and inclusion statement

Our team members’ positionalities were a consideration in this review to ensure data was analysed and reported with varied lens’ and to ensure Indigenous ways of knowing, being, and doing[28] were at the forefront of this review. Our team includes experienced and early career researchers working in the areas of musculoskeletal health (CW, IL, SD, EA, AT, JL) and Australian Aboriginal and Torres Strait Islander health (VM, EW, IL, JL). Three members of the team are Australian Aboriginal researchers (VM, EW, JL). The research team includes clinical physiotherapists (IL, AT, JL) working in multiple settings: public hospital, and private and Indigenous primary health services. Our team has an interest in the review findings to improve the health and wellbeing of Indigenous peoples and to inform future musculoskeletal health programs, including in rural health care settings.

### Search Strategy

A systematic search strategy was developed by our research team with assistance from a university librarian. The search strategy consisted of combining three key concepts: Indigenous peoples, musculoskeletal conditions, and health programs. Electronic databases (Medline, Embase, CINAHL, Proquest Central, Scopus) were searched from January 1993 to November 2023. Additional searches were also performed in Australian Indigenous Health Infonet bibliography (http://www.healthinfonet.ecu.edu.au) and Google Scholar (first five pages) to identify any relevant studies not identified from the database search. The reference list and citation list of all included articles were searched for additional studies. A list of included studies was sent to experts in the field, known to the research team, to provide advice on any known omissions. Results were downloaded to EndNote (Clarivate) and automatically deduplicated. The studies were then uploaded to Covidence (Veritas Health Information, Melbourne, Australia) for screening and had further automatic deduplication. Complete details of the search terms are provided in Supplemental Table 2.

An eligibility criterion for included studies was developed by our research team and included health programs for Indigenous adults of the CANZUS region with musculoskeletal pain conditions or education programs for health staff working with Indigenous people with musculoskeletal pain conditions (see Supplemental Table 3 for full criterion). The study titles and abstracts were screened by two independent reviewers. Full text screening was conducted by two independent reviewers. Instances where there were differences in reviewer opinions were resolved by discussion and consensus between reviewers or a decision by a third reviewer. Reasons for exclusions were recorded.

### Data extraction

Data was extracted from included articles by one author and reviewed by a second author. Any conflicts were discussed, and consensus achieved with discussion of the two reviewers and other members of the research team. Data was extracted using an agreed template to capture study characteristics (e.g. year, location, design, outcomes), participant demographics (cultural identification, health condition), program settings/locations and the cultural considerations of the program development and delivery.

### Data charting and synthesis

Studies were grouped during the data extraction phase into two categories for synthesis: programs for Indigenous peoples and programs for health staff working with Indigenous peoples. Descriptive statistics were used to synthesise some characteristics across studies, for example country, musculoskeletal condition. The cultural considerations for the development and delivery of the programs and the study outcomes were synthesised descriptively.

### Methodological and Cultural Quality Appraisal

Study design was appraised from two perspectives (methodological quality and cultural considerations). The Mixed Methods Appraisal Tool (MMAT),[29] was used by two authors independently to assess methodological quality. A third opinion was sought if consensus was unable to be reached. The MMAT has different criteria dependent on the study type, has content validity and moderate to good interrater reliability.[30] Cultural considerations in the study design, delivery, and reporting were assessed with two methods. The Aboriginal and Torres Strait Islander QAT[31] was used for the Australian studies. The content validity and reliability of this tool have been assessed and reported elsewhere.[21] The CONSIDER statement checklist[20] was used to appraise all studies. The statement was developed by Indigenous researchers from countries included in our review. As the original purpose of the statement was to establish principles to guide the development of Indigenous health research there is no information on the reliability and validity of this checklist to assess studies. The team discussed, trialled, and decided on a scoring criterion, with ‘yes’ scored when the principle was documented and ‘no’ scored when the principle was not documented in the study.[22] Two methods were chosen as there is currently no validated tool to assess cultural considerations of studies in the included countries. The research team was interested in the comparability of both methods, which will be discussed in a future paper. Data was extracted by one author and reviewed by a second author. A third opinion was sought if consensus was unable to be achieved.

## RESULTS

Database searches identified 5472 unique studies, and four studies were identified from additional search strategies. Following title and abstract screening, 106 full article texts were reviewed, and 23 studies were eligible for inclusion (see PRISMA flowchart - Figure 1).

**Figure 1.**
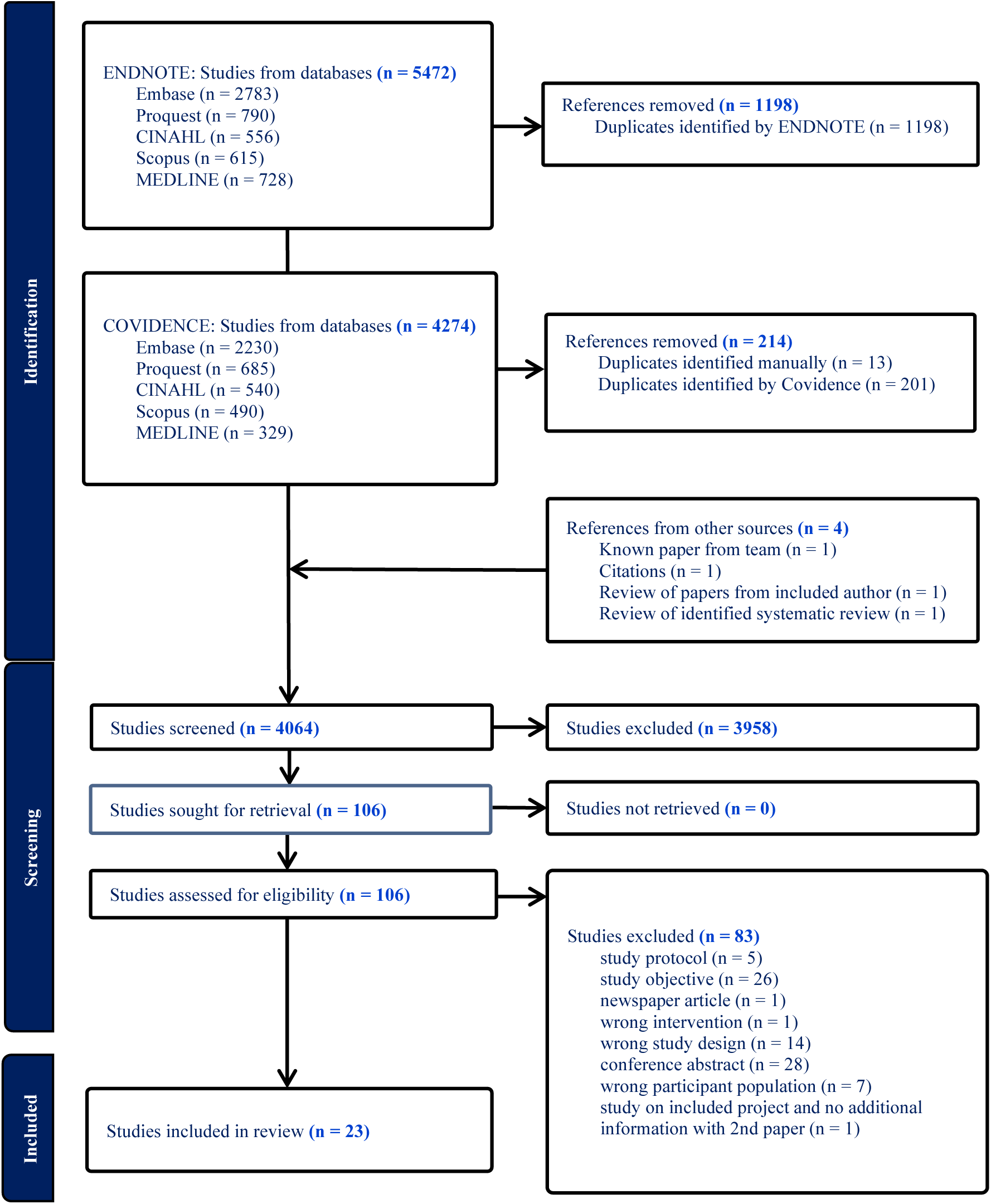
Preferred Reporting Items for Systematic and Meta-Analyses flow diagram for scoping review

There were seven qualitative, three quantitative and 13 mixed methods studies. There were 16 studies reporting programs delivered to Indigenous peoples (including four studies about different aspects of two programs,[32–35]) and seven studies reporting programs targeting health staff (see Table 1). Strategies utilised in each study were identified and summarised in Table 2. There were six strategies in the health programs for Indigenous peoples and three strategies in the health staff programs.

**Table 1.**
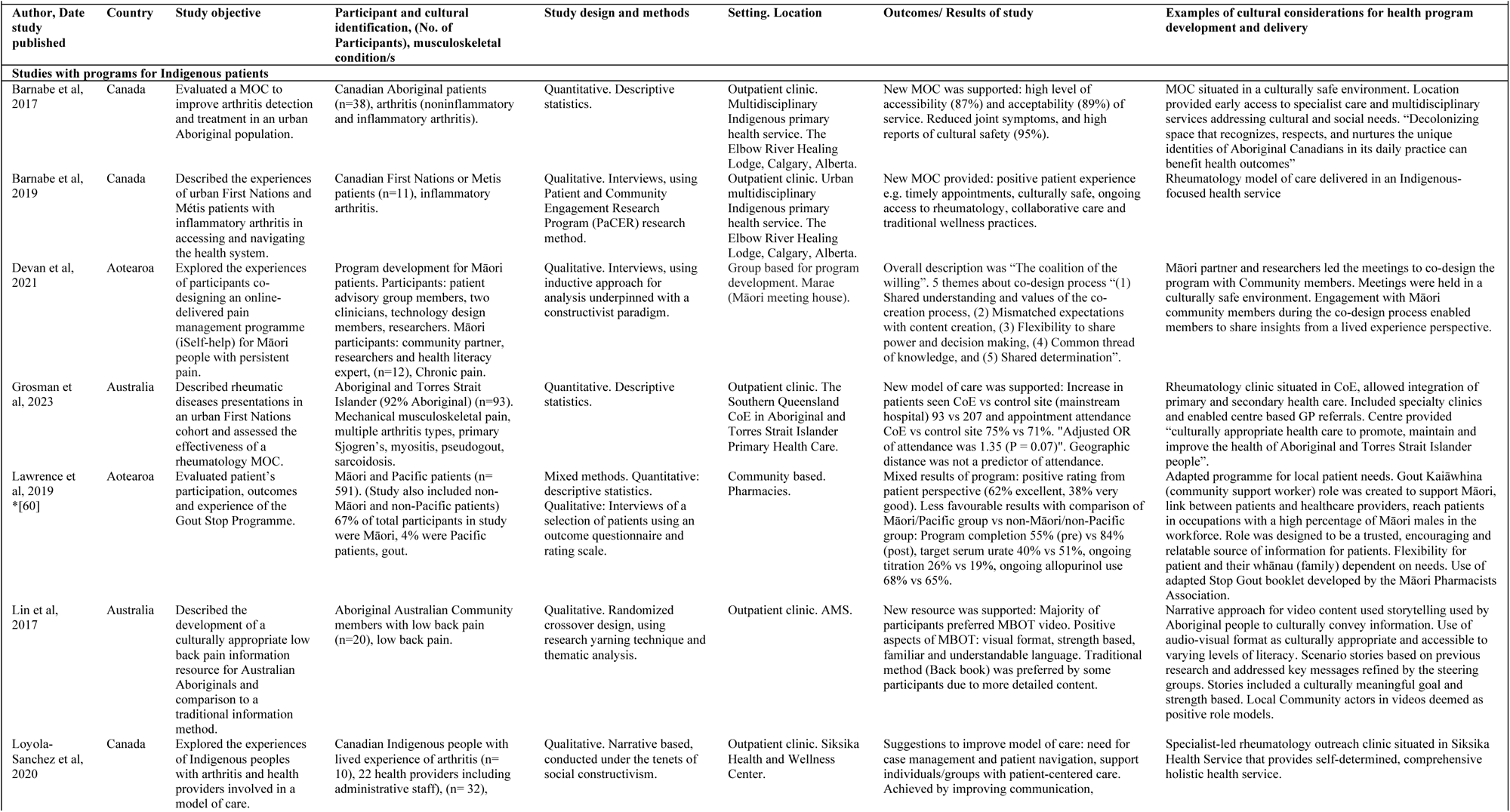
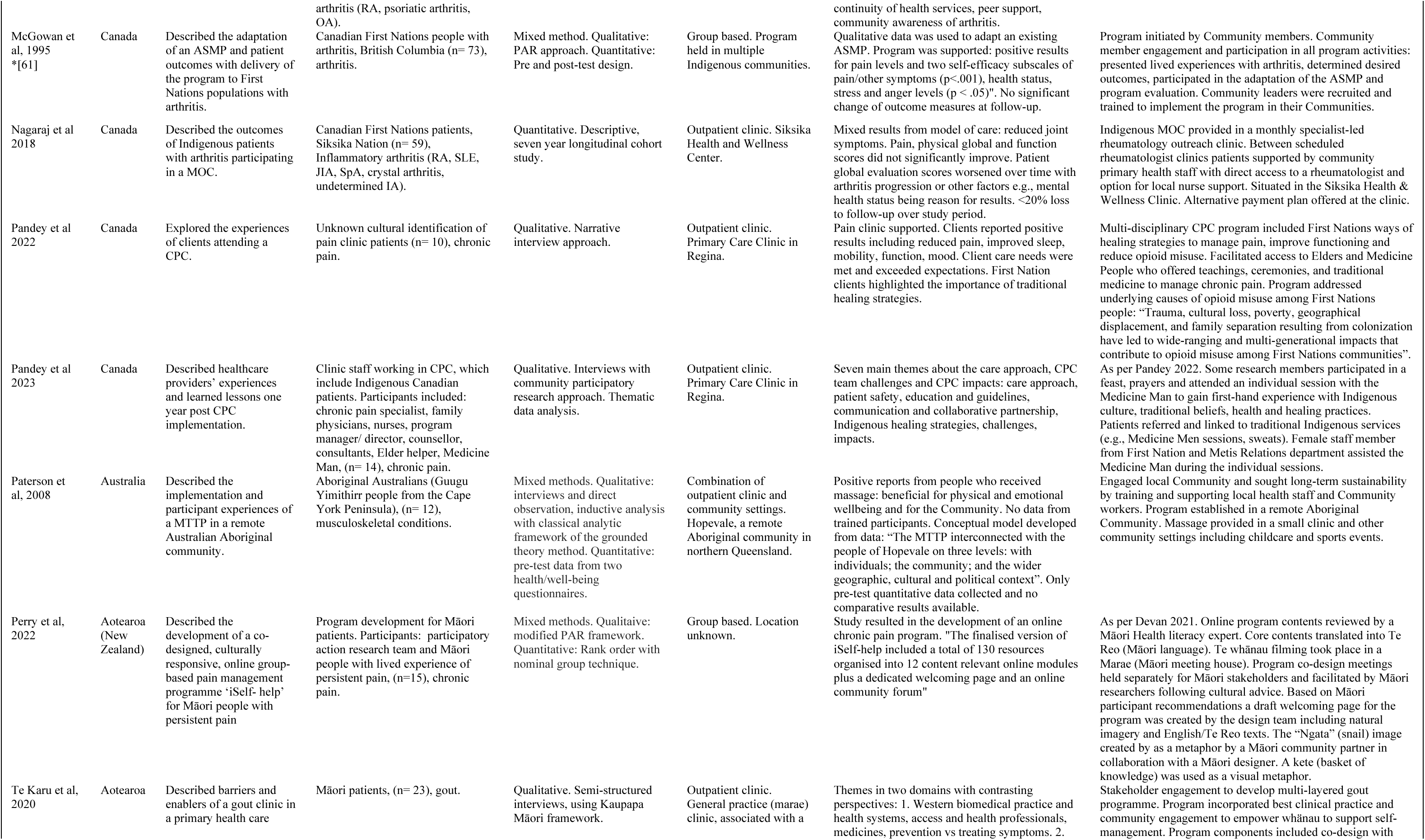
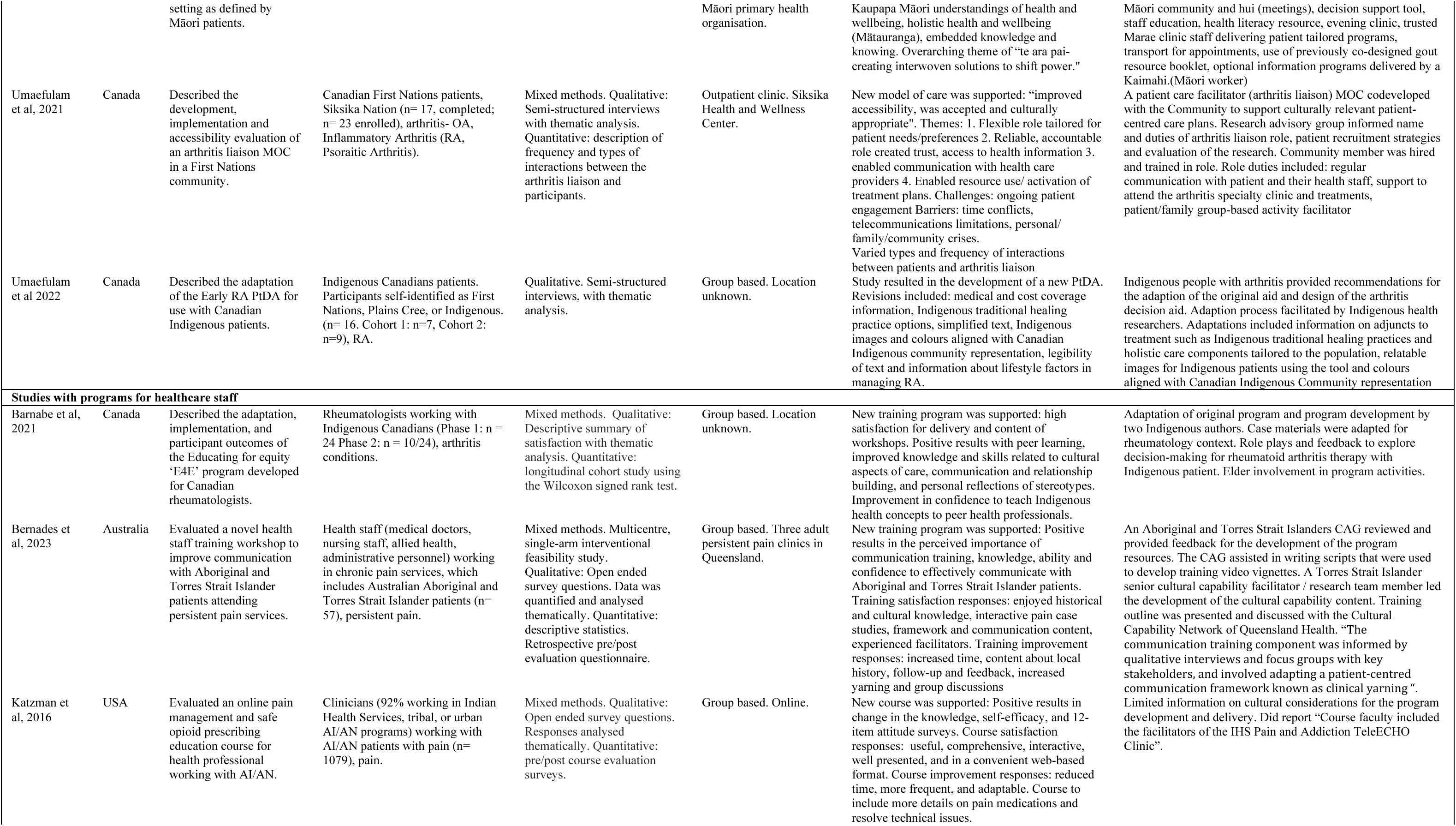
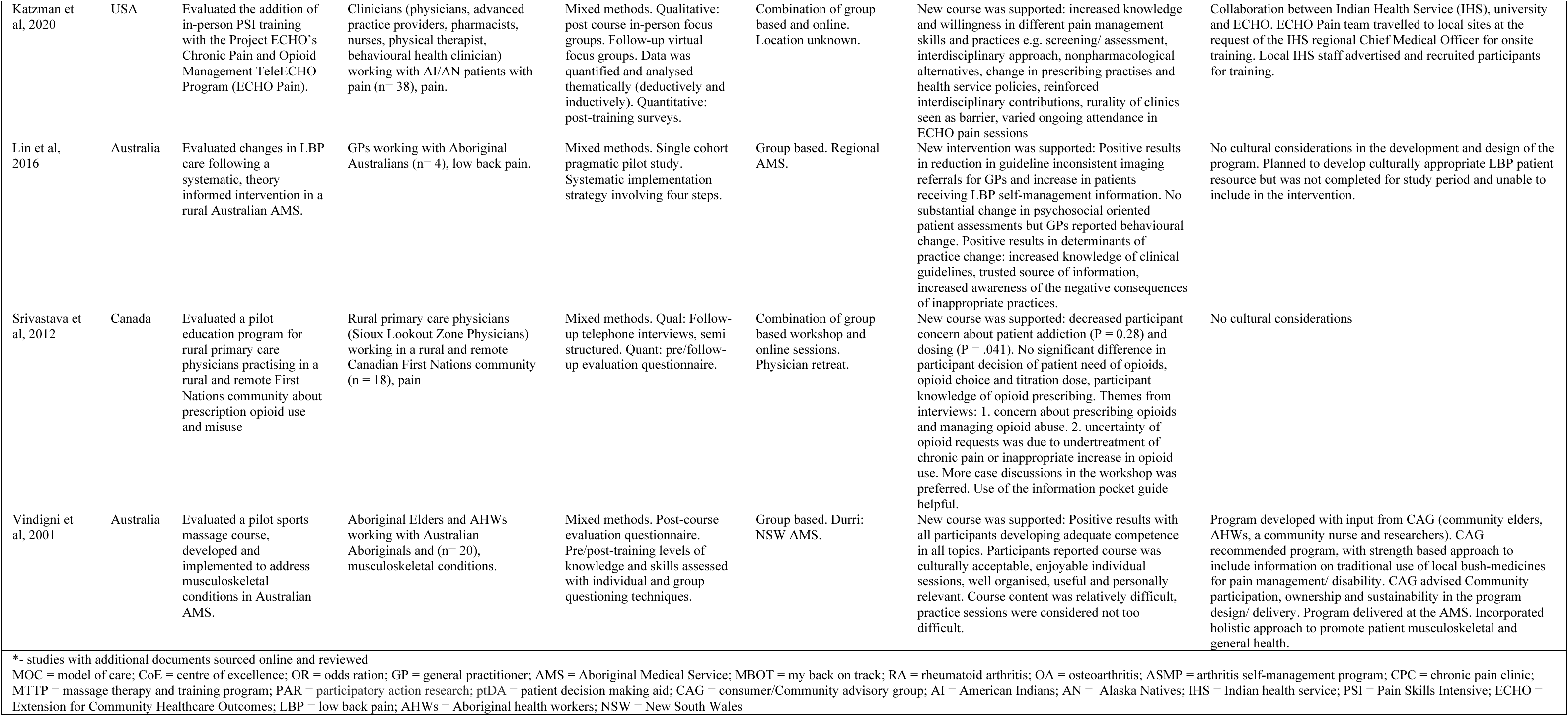
Characteristics of the included studies.

**Table 2.** Program strategy type.

| Table 2. Program strategy type |  |  |  |  |  |  |  |  |  |
| --- | --- | --- | --- | --- | --- | --- | --- | --- | --- |
| Lead Author, Date | Indigenous patient program strategies |  |  |  |  |  | Health staff training program strategies |  |  |
|  | Location | Model of Care | Indigenous Support Person | Developed Culturally adapted program/resource | Developed New program/resource | Utilised existing resource | Skills based EBP training | Culturally informed training | Training with cultural components |
| **Studies with programs for Indigenous patients** |  |  |  |  |  |  |  |  |  |
| Barnabe, 2017 | ✓ | ✓ |  |  |  |  |  |  |  |
| Barnabe, 2019 | ✓ | ✓ |  |  |  |  |  |  |  |
| Devan 2021 / Perry, 2022 |  |  |  | ✓ |  |  |  |  |  |
| Grosman, 2023 | ✓ | ✓ |  |  |  |  |  |  |  |
| Paterson, 2008 | ✓ |  |  |  |  |  |  |  |  |
| Lawrence, 2019 *[60] |  |  | ✓ |  |  | ✓ |  |  |  |
| Lin, 2017 |  |  |  |  | ✓ |  |  |  |  |
| Loyola Sanchez, 2020 | ✓ | ✓ |  |  |  |  |  |  |  |
| McGowan, 1995 *[61] |  |  |  | ✓ |  |  |  |  |  |
| Nagaraj, 2018 | ✓ | ✓ |  |  |  |  |  |  |  |
| Pandey 2022 |  |  |  |  | ✓ |  |  |  |  |
| Pandey 2023 |  |  |  |  | ✓ |  |  |  |  |
| Te Karu, 2020 | ✓ | ✓ |  |  |  | ✓ |  |  |  |
| Umaefulam 2021 |  | ✓ | ✓ |  |  |  |  |  |  |
| Umaefulam 2022 |  |  |  | ✓ |  |  |  |  |  |
| **Studies with programs for healthcare staff** |  |  |  |  |  |  |  |  |  |
| Barnabe, 2021 |  |  |  |  |  |  |  | ✓ |  |
| Bernades, 2023 |  |  |  |  |  |  |  | ✓ |  |
| Katzman, 2016 |  |  |  |  |  |  | ✓ |  |  |
| Katzman, 2020 |  |  |  |  |  |  | ✓ |  |  |
| Lin, 2016 |  |  |  |  |  |  | ✓ |  |  |
| Srivastava et al, 2012 |  |  |  |  |  |  | ✓ |  |  |
| Vindigni, 2004 |  |  |  |  |  |  |  |  | ✓ |
| *-. studies with additional documents sourced online and reviewed<br>MSK = musculoskeletal; EBP = evidence based practice |  |  |  |  |  |  |  |  |  |

### Description of included studies

#### Programs for Indigenous patients

Of the 16 studies that were patient interventions, nine were from Canada.[35–43] Three studies[44–46] were from Australia and four studies[32, 33, 47, 48] from Aotearoa. Musculoskeletal conditions included were arthritis (n = 5),[36, 38, 39, 42, 44] chronic pain of any origin (n = 4),[33, 35, 41, 49] gout (n = 2),[47, 48] multiple inflammatory arthritis conditions (n = 2),[37, 40] low back pain (n = 1),[45] rheumatoid arthritis (RA) (n = 1),[43] and general musculoskeletal conditions (n = 1).[46]

Participants were primarily Indigenous peoples participating in the health program (n = 8).[36, 37, 39, 43, 44, 46, 48, 50] The cultural identification of participants in a chronic pain clinic was not specified in one study.[41] Māori patients were the primary (67%) participant group in one study.[47]

Participants in one study were a combination of Indigenous peoples and health staff[38] and another study’s participants were health staff implementing a program developed for patients with chronic pain (n = 1).[35] The remaining four studies described the development of a program or resources. Of these studies the participants included Indigenous people with RA (n = 1),[43] Indigenous Community members (n = 1),[45] and a combination of research team, including Indigenous members and Indigenous Community members (two studies about one program).[33, 49]

The most common setting was an outpatient/primary health care clinic with seven located in Indigenous primary health services[36–38, 40, 42, 44, 48] and two did not specify the clinic type.[35, 41] Two studies were based in Indigenous Communities,[39, 46] and one in community-based pharmacies.[47] For the program/resource development studies two studies conducted participant interviews in an Indigenous primary health setting[45, 49] and two studies did not specify the location.[33, 43]

The types of programs included culturally informed clinics for different conditions: gout (n = 1),[48] chronic pain (n = 2)[35, 41] and delivering a rheumatology model of care (n = 5).[36–38, 40, 44] An Indigenous support worker was incorporated into a tailored model of care.[42] There was one study with a community-based gout support worker[47] and one study described a massage program situated in a remote Aboriginal Community[46]. Culturally adapted health programs were delivered in person[39] and online.[33, 49] The two developed resources included educational videos for low back pain management[45] and a RA patient decision aide.[42]

All studies described cultural considerations in the development of the health programs e.g., the involvement of a Community advisory group (CAG) and Indigenous researchers (see Table 1). Most studies were situated in Indigenous Communities or primary health care settings,[33, 36–38, 40, 42, 44, 48, 49] and others used health information resources designed specifically for Indigenous Communities.[45, 47, 48] Only a few incorporated Indigenous cultural practices such as traditional healing practices and medicine people.[37, 41, 43]

The overall outcomes of the programs were positive. In qualitative studies the participants reported a positive experience[37, 42, 47] and the programs were accessible, acceptable and culturally safe. In the quantitative studies there was improvement in participant objective measures such as joint symptoms,[36, 40] pain and self-efficacy[39] and improvement in service delivery measures (e.g. patient attendance).[44] One study that measured comparative participant data found a less optimal outcome for Māori participants to program completion and gout medication management.[47] An Indigenous health information resource was also received positively from the participant perspective.[51]

#### Programs for health staff education

Of the seven education programs for health staff, three were from Australia,[52–54] two from the United States of America[55, 56] and two from Canada.[57, 58] The musculoskeletal conditions included were general musculoskeletal conditions (n = 1),[54] low back pain (n = 1),[53] chronic pain (n = 1),[52] arthritis conditions (n = 1)[57] and non-specified pain (n = 3).[55, 56, 58] Participants included rheumatologists (n = 1),[57] Australian Aboriginal health workers and Elders (n = 1),[54] general practitioners (n = 1)[53, 58] and varied health professionals (n = 3).[52, 55, 56] Settings included Indigenous primary health services (n = 2),[53, 54] three persistent pain clinics (n = 1),[52] online training (n = 1),[55] and a combination of in-person group based training and online sessions (n = 2).[56, 58] One study did not specify the program setting.[57]

Four studies included musculoskeletal related evidence-based education for health professionals working with Indigenous people with musculoskeletal pain conditions.[53, 55, 56, 58] This included training on safe opioid prescribing and chronic pain management information,[55, 56, 58] and low back pain management.[53] Three studies had cultural components in the training. This included a massage program with traditional healing practices,[54] the use of a culturally informed communication technique called clinical yarning,[52, 59] and participating in cultural capability training.[52]

Four of the seven programs for health staff included cultural considerations in the design and delivery of their programs, such as the involvement of Indigenous researchers or a CAG in the design of the training program.[52, 54, 57] Health programs were also delivered by Indigenous co-facilitators and incorporated case studies of Indigenous patients with musculoskeletal conditions into the training.[52, 57] The remaining three studies delivered skills based pain and opioid management training. Although the participants were identified as health professionals working with Indigenous patients there was none or very limited reporting of cultural aspects being considered in the design and delivery of the education programs.[53, 55, 58]

The outcome of the programs was varied between the studies, but the results were overall positive. Participant’s knowledge on the program’s topic (e.g. appropriate back pain care, communication technique, opioid management) was increased. The two programs that focussed on improving culturally responsive practices also resulted in an improvement in participant’s confidence to work with Indigenous patients, provide peer learning for colleagues on the topic and the ability to self-reflect on personal beliefs and practices.

### Quality Appraisal

#### Study Design

There were 14 studies that scored ‘yes’ to all five criteria (see Table 3) of the MMAT.[30] Two studies scored no/can’t tell on the majority of the criterion.[46, 47] Mixed method studies were more likely to have design flaws. Only 25% of the mixed methods studies scored yes to all criterion and eight of the twelve mixed methods studies did not meet the final criteria ‘(5): Do the different components of the study adhere to the quality criteria of each tradition of the methods involved?’

**Table 3.** Mixed Methods Appraisal Tool (MMAT) scores [30].

| Table 3. Mixed Methods Appraisal Tool (MMAT) scores [30] |  |  |  |  |  |  |  |  |
| --- | --- | --- | --- | --- | --- | --- | --- | --- |
| Lead Author, Date | Category of study design | S1 | S2 | 1 | 2 | 3 | 4 | 5 |
| **Studies with programs for Indigenous patients** |  |  |  |  |  |  |  |  |
| Barnabe, 2019 | Qualitative | yes | yes | yes | yes | yes | yes | yes |
| Devan, 2021 | Qualitative | yes | yes | yes | yes | yes | yes | yes |
| Lin, 2017 | Qualitative | yes | yes | yes | yes | yes | yes | yes |
| Loyola Sanchez, 2020 | Qualitative | yes | yes | yes | yes | yes | yes | yes |
| Pandey, 2022 | Qualitative | yes | yes | yes | yes | yes | yes | yes |
| Pandey, 2023 | Qualitative | yes | yes | yes | yes | yes | yes | yes |
| Te Karu, 2020 | Qualitative | yes | yes | yes | yes | yes | yes | yes |
| Umafulam, 2022 | Qualitative | yes | yes | yes | yes | yes | yes | yes |
| Barnabe, 2017 | Quantitative descriptive | yes | yes | yes | yes | yes | CT | yes |
| Grosman, 2023 | Quantitative descriptive | yes | yes | yes | yes | yes | yes | yes |
| Nagaraj, 2018 | Quantitative descriptive | yes | yes | yes | yes | yes | yes | yes |
| Lawrance, 2019 *[60] | Mixed methods | yes | yes | CT | CT | CT | CT | no |
| Macgowan, 1995 *[61] | Mixed methods | yes | yes | yes | yes | yes | no | no |
| Paterson, 2008 | Mixed methods | yes | yes | no | CT | CT | yes | no |
| Perry, 2022 | Mixed methods | yes | yes | yes | yes | yes | yes | yes |
| Umafulam, 2021 | Mixed methods | yes | yes | no | yes | no | yes | no |
| **Studies with programs for healthcare staff** |  |  |  |  |  |  |  |  |
| Barnabe, 2021 | Mixed methods | yes | yes | yes | yes | yes | yes | yes |
| Bernades, 2023 | Mixed methods | yes | yes | yes | yes | yes | yes | yes |
| Katzman, 2016 | Mixed methods | yes | yes | yes | yes | no | CT | no |
| Katzman, 2020 | Mixed methods | yes | yes | yes | yes | yes | yes | CT |
| Lin, 2016 | Mixed methods | yes | yes | yes | yes | yes | yes | yes |
| Srivastava, 2012 | Mixed methods | yes | yes | no | yes | yes | yes | no |
| Vindigni, 2004 | Mixed methods | yes | yes | no | yes | CT | yes | no |
| CT- can't tell, *- studies with additional documents sourced online and reviewed<br>S1: Are there clear research questions?<br>S2: Do the collected data allow to address the research questions?<br>Criteria 1-5<br>Qualitative:<br>1. Is the qualitative approach appropriate to answer the research question?<br>2. Are the qualitative data collection methods adequate to address the research question?<br>3. Are the findings adequately derived from the data?<br>4. Is the interpretation of results sufficiently substantiated by data?<br>5. Is there coherence between qualitative data sources, collection, analysis and interpretation?<br>Quantitative descriptive<br>1. Is the sampling strategy relevant to address the research question?<br>2. Is the sample representative of the target population?<br>3. Are the measurements appropriate?<br>4. Is the risk of nonresponse bias low?<br>5. Is the statistical analysis appropriate to answer the research question?<br>Mixed Methods<br>1. Is there an adequate rationale for using a mixed methods design to address the research question?<br>2. Are the different components of the study effectively integrated to answer the research question?<br>3. Are the outputs of the integration of qualitative and quantitative components adequately interpreted?<br>4. Are divergences and inconsistencies between quantitative and qualitative results adequately addressed?<br>5. Do the different components of the study adhere to the quality criteria of each tradition of the methods involved? |  |  |  |  |  |  |  |  |

#### Cultural appraisal

All studies were reviewed using the CONSIDER checklist[20] (see Supplemental Table 4). Except for one study,[58] all studies reported some cultural considerations in the study design and implementation (see Table 4). The three most common criteria met were 4: prioritisation of Indigenous people, organisations, and empirical evidence, 9: research methodology incorporated Indigenous worldviews, 17: dissemination to support Indigenous advancement. The criteria that were not reported in all studies were 10: participation resulting in consent for use of collected data for future analysis, 12: participation resulting in agreement with data storage and disposal and 3: governance with research agreements to protect Indigenous intellectual property and knowledge.

**Table 4.**
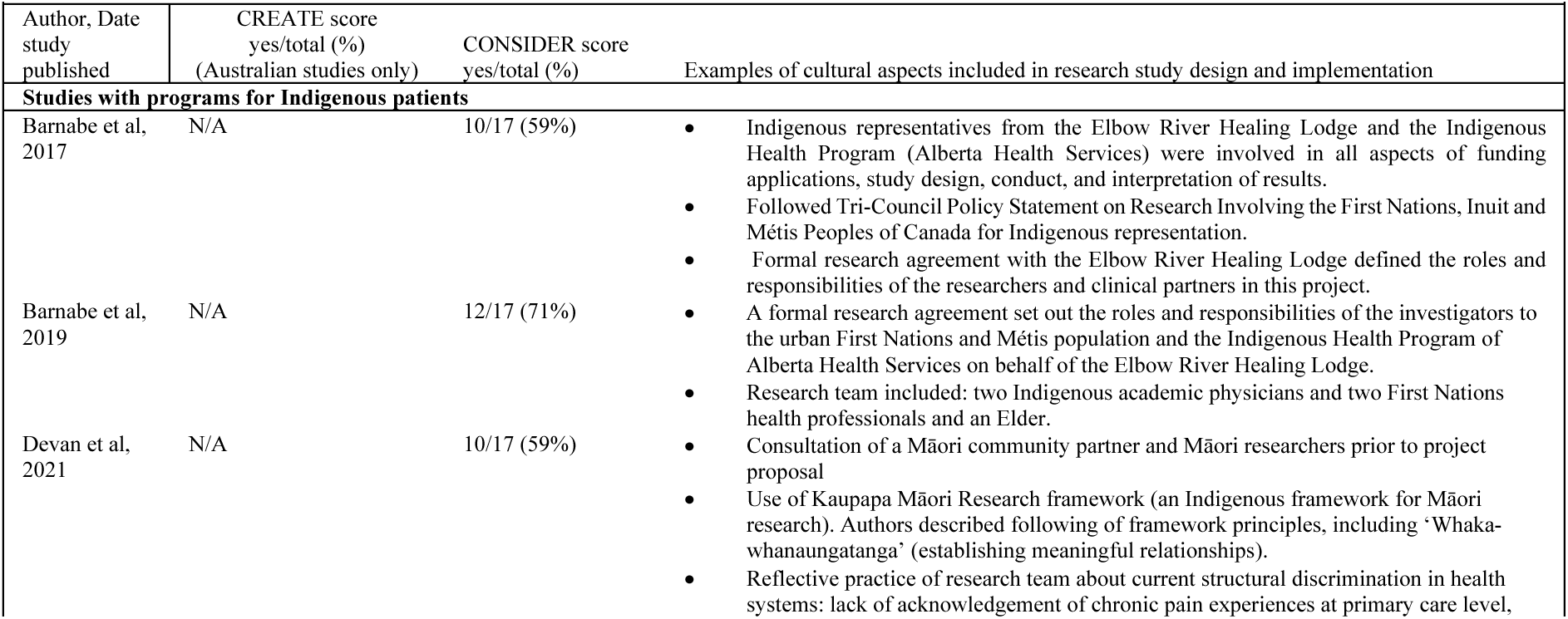
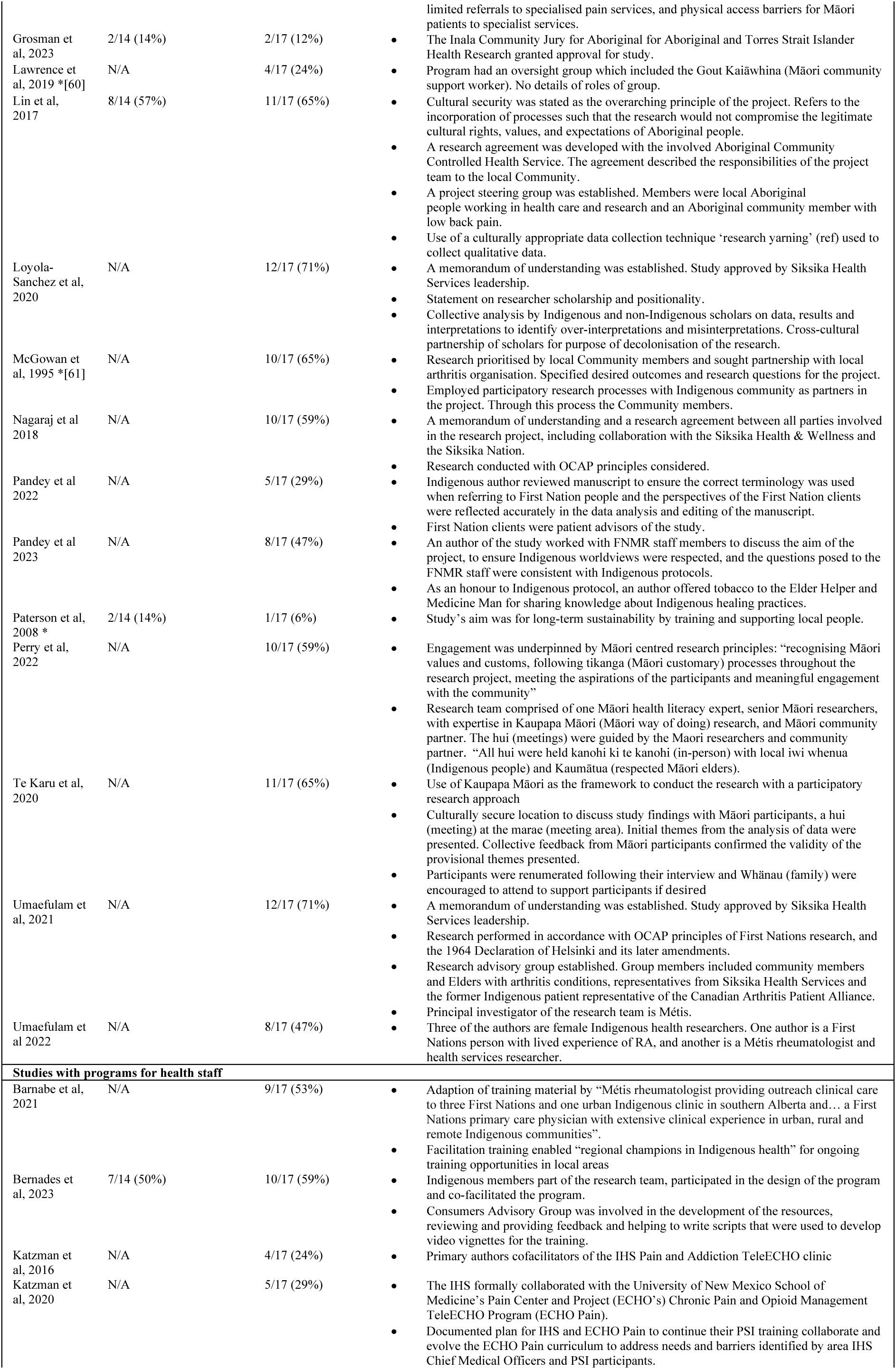
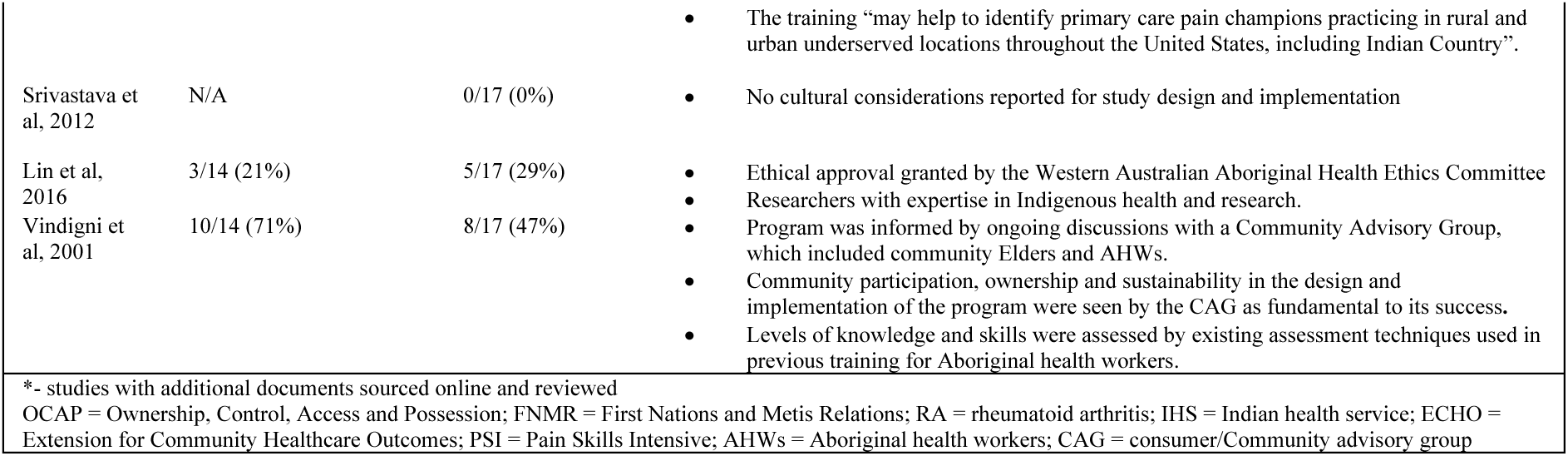
Cultural consideration for each research study and overall score for Aboriginal and Torres Strait Islander QAT and CONSIDER appraisal review.

Six Australian studies were also reviewed using the Aboriginal and Torres Strait Islander QAT[31] (see Supplemental Table 5). Three studies scored ≥50% and there was no observable pattern with the scores when study design or date of the study was compared. Three Aboriginal and Torres Strait Islander QAT criteria were met or partially met by most studies, including 3: Indigenous leadership, 10: strength-based approach, 12: benefit to Community. The Aboriginal and Torres Strait Islander QAT criteria that was not fully met by any studies was 6 and 7: intellectual and cultural property protection and ownership and 8: data sovereignty. There was strong agreement between Aboriginal and Torres Strait Islander QAT and CONSIDER assessment findings in Australian studies, which will be reported in a future paper.

## DISCUSSION

The purpose of our review was to provide a comprehensive summary of the features of the health program’s development and delivery, program outcomes and how culture was considered in each study. The results provide clear guidance for clinicians, Indigenous Communities and researchers about program designs that may be beneficial within their local context. Figure 2 was created to provide an accessible method to communicate the findings of the scoping review and the recommended future directions of musculoskeletal pain research with Indigenous Communities.

**Figure 2:**
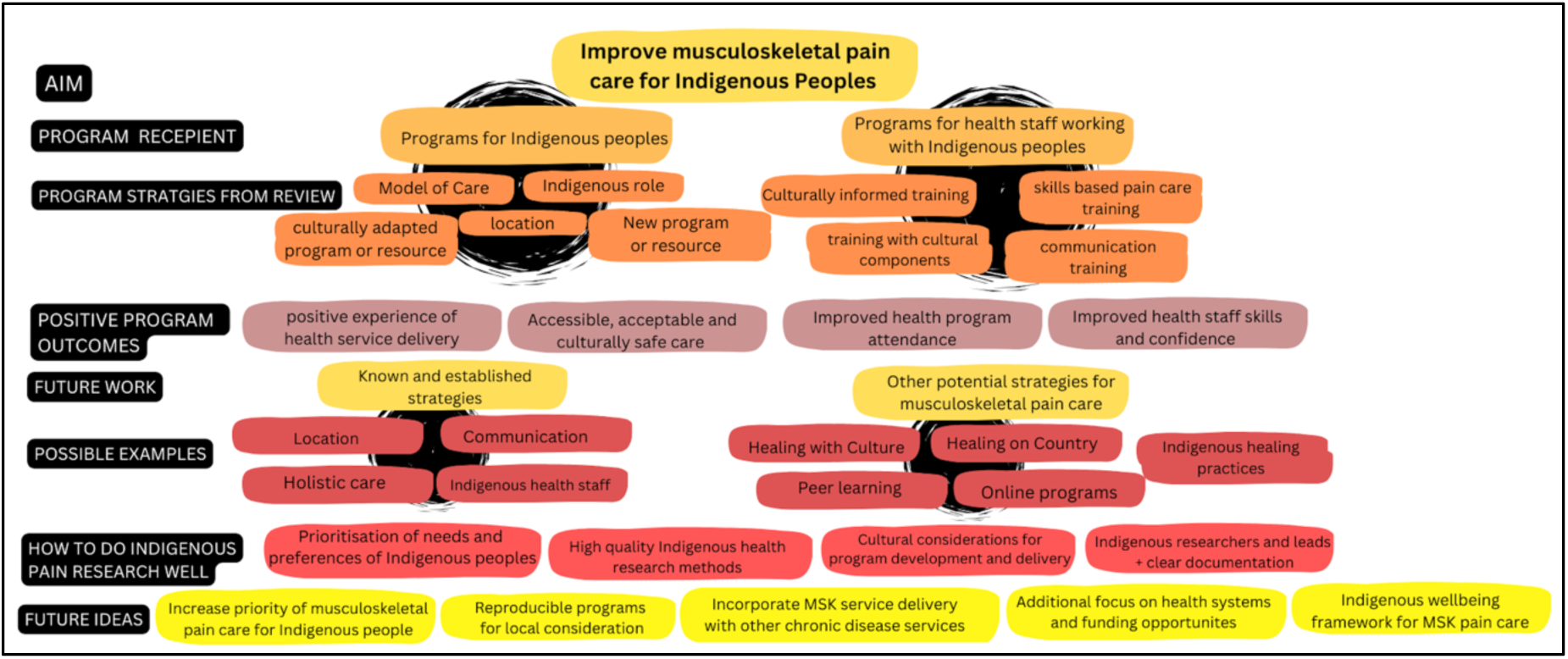
Scoping review summary and recommendation for future research in health programs for Indigenous people with musculoskeletal pain conditions

Our review summarises 23 studies, including 16 targeting Indigenous peoples with musculoskeletal pain and seven targeting health staff. The musculoskeletal pain programs identified several strategies aimed at both patients and health staff, focussed on Indigenous strength-based approaches. The strategies included offering care within an Indigenous health service,[36–38, 40, 42, 44, 48] using Indigenous support persons,[42, 47] and developing educational videos with Indigenous actors.[33, 45, 52] Overall, the studies had moderate-to-high methodological quality. From a cultural perspective, the quality of the studies was moderate.

### Implications of current evidence

The most common strategy was providing the care in a culturally secure location, such as Indigenous health services[36–38, 40, 42, 44, 48] and resulted in positive outcomes, including improved patient experiences,[36, 37, 42] and increased appointment attendance.[44] Indigenous health services are reported to be preferred health care locations for Indigenous peoples because they offer affordable care, multidisciplinary options, and are governed by Indigenous people (self-determination).[62] The use of traditional healing practices and medicines for pain management[37, 41, 54] was also identified as a preferred pain management strategy by Indigenous peoples. This method privileges Indigenous ways and aligns with previous research,[63, 64] policy recommendations[65] and international human rights.[66] Strength-based approaches have been shown to emphasise the positive contributions of Indigenous Communities and practices in managing health issues and countering negative stereotypes. Types of approaches include individual empowerment, cultural protective factors, and holistic care,[14, 67] and can foster the flourishing of Indigenous peoples[68] and the active participation of Indigenous Communities in research processes.[69]

Included studies also incorporated holistic care, including the management of other chronic health conditions into musculoskeletal pain programs.[41, 54] A holistic approach is integral to Indigenous ways of doing[28] and health practices.[70] Health conditions that are often co-morbid with musculoskeletal pain such as diabetes and cardiovascular disease, share common risk factors (e.g. obesity and smoking) and management strategies (e.g. exercise). Future programs for musculoskeletal pain could be developed or integrated with other chronic disease programs in Indigenous health services. This holistic approach avoids siloing health conditions and may improve sustainability. Two studies demonstrated examples of successful frameworks and models of care[36, 48] that guided holistic health in musculoskeletal pain care. Other frameworks, such as The Good Spirit, Good Life[71] framework, developed with and for Australian Aboriginal Elders exemplifies how holistic models of wellbeing, if developed for musculoskeletal pain, could be used to guide care and development of an Indigenous wellbeing framework for musculoskeletal care.

Included studies described the processes involved in developing a health program or resource[33, 43, 51] and evaluated the experiences, including identified challenges, of research team members, health staff and Community members involved in delivering the health programs.[32, 35, 38, 42] The interactions at the research interface between Western and Indigenous research methods[72] is an important consideration for researchers working with Indigenous Communities. The additional information from introspective studies can provide valuable information to inform future work in this area and ensure quality research practices.

A common attribute of studies scoring higher in the cultural appraisal was evidence of leadership from researchers who had collaborated on multiple studies in this area over time.[36, 45] These studies typically incorporated more cultural features such as embedding cultural governance structures to research projects and establishing research agreements between research institutions and Indigenous health services. The correlation between strong leadership and higher cultural quality may highlight the responsibilities of researchers working with local Indigenous Communities and the value of sustained engagement by research teams in this area who demonstrate greater cultural methodological expertise. The ongoing work of these teams appears to have also driven an increase in research output in this field over time.

The studies with high cultural quality scores also had two important layers of Indigenous peoples’ involvement, authorship and inclusion of a CAG. Indigenous scholarship has been demonstrated to have a positive correlation with the quality of papers from a cultural perspective and has been identified as a key aspect of cultural competency in research.[73] The inclusion of a CAG in research teams was documented in four studies.[33, 43, 45, 49] The co-design of studies with Indigenous Community members can drive the objective and anticipated outcomes of the projects, ensures the design and development of programs or resources reflects the needs and preferences of the local Indigenous Communities and can minimise the ‘retrofitting’ of mainstream programs for selected cultural groups.[74]

### Gaps and Future Directions

The ability to ascertain Indigenous scholarship and the contribution of a CAG was not always clear in the included studies. Clear documentation of Indigenous peoples involvement in the projects allows readers the knowledge of who has been participated in the research project and in what capacity, and aligns with recommendations from Indigenous scholars[75] and appraisal tools.[20, 21] Another key area identified was that most studies did not document the research project’s Indigenous data sovereignty processes or governance structures, including data storage and protection of Indigenous intellectual and cultural property. Data sovereignty is of increased focus for Indigenous academics[76, 77] and is an identified right of Indigenous peoples supported by the United Nations declaration on the rights of Indigenous peoples.[78]

In addition to describing musculoskeletal pain programs and services, studies identified future directions for musculoskeletal pain research with Indigenous communities. These priorities included health system processes and the funding arrangements needed to achieve sustainable health equity[74], determining if training health staff results in improved clinical outcomes for Indigenous patients[52, 79] and the sustainability of these programs, including the development of local Community champions.[56, 79] While health staff programs demonstrated an improvement in health staff skills and confidence in pain care, they lacked the inclusion of cultural learning objectives.[53, 55, 56, 58] Combining musculoskeletal pain training with culturally informed approaches, such as antiracism[80–82] and cultural humility[83] education could potentially enhance the effectiveness of these programs.

The included studies were primarily from Canada, Australia and Aotearoa. There was a notable absence of patient programs for Native Americans with previous pain research from the USA focussed on health disparity within a wider population, including African Americans and immigrants.[84–86] Previous pain research from the USA has focussed on health disparity with a wider inclusion population, including African Americans and immigrants. Research with a focus on working with Indigenous peoples of the USA is a key priority in musculoskeletal pain care projects.

Furthermore, two other recommendations for future research included incorporating peer learning and support into health programs[38, 41] and clearly documenting program components in publications[45, 52] to enable translation of effective health programs into clinical settings within the local context of Indigenous Communities.

#### Strengths and Limitations

Our research team made the decision to only include studies from the CANZUS region. The countries included were chosen because they share similar English colonial histories as a primary upstream health determinant, are high income countries, and Indigenous peoples are minority populations with significant health disparities between Indigenous and non-Indigenous peoples. We recognise the diversity of Indigenous peoples globally. Future studies, with a boarder inclusion of Indigenous groups, could examine musculoskeletal pain care amongst other Indigenous peoples in different historical and social contexts including the similarities and differences with our findings. Collaborative reviews undertaken with Indigenous researchers from each country would ensure the voices of the included Indigenous groups were privileged.

A strength of our study is the use of cultural methodological appraisal tools, in addition to traditional appraisal tools. In doing so, we provide a rigorous analysis from two different but equally important perspectives on how research is conducted with Indigenous peoples. We used rigorous review methods including comprehensive article search and duplicate screening and data extraction by multiple authors. The inclusion of varied perspectives-Indigenous and non-Indigenous, clinicians and academics in this review process ensured a thorough and considered assessment was conducted.

Potential limitations include that we did not have representation of Indigenous researchers from countries other than Australia. The inclusion of researchers from other Indigenous groups from CANZUS and CAG members[87] in the team would have provided an additional lens’ and gained a wider perspective and interpretation of the studies. We only included literature published in peer reviewed journals which may have omitted other useful programs reported elsewhere, including in grey literature [88, 89] or in the media,[90] however this literature may have not included sufficient detail to appraise cultural and research quality. That we identified 23 studies in total highlights the limited amount of evidence in this area. Given the diversity, local considerations and priorities between Indigenous groups between countries, Indigenous nations and tribal groups, this body of research is likely to evolve considerably as this body of research develops over time.

## Conclusion

Targeted programs for musculoskeletal pain have been developed in the CANZUS region. There are a number of successful cultural strategies, including holistic models of care and Indigenous led training programs for health staff, that can provide a starting point for Indigenous Communities, clinicians and Indigenous and mainstream health organisations to improve the health and wellbeing of Indigenous peoples with musculoskeletal pain. There are also opportunities to improve the processes involved in conducting research with Indigenous Communities, a vital addition to our research practices[91] if we are to continue to work meaningfully with Indigenous peoples with musculoskeletal pain and achieve the universal goal to live a good life.

## Data Availability

All data produced in the present work are contained in the manuscript

## Author contributions

JL, CMW, IL, SRED designed the study. JL ran the searches. JL, SRED, ESA and AT conducted the study screening and data extraction. JL, IL, CMW, SRED, ESA conducted the data analysis, interpretation and synthesis. JL wrote the first draft of the manuscript, and all authors provided critical feedback. All authors contributed to and provided feedback on drafts, read and approved the final version of the manuscript, and were responsible for the decision to submit for publication.

## Acknowledgements

We acknowledge the University of Sydney librarian, for her assistance with the search strategy. JL was supported by the CRE-STRIDE scholarship funded by the National Medical Research Council of Australia. CMW receives funding for salary from the National Medical Research Council of Australia.

---

**Supplemental Table 1.**
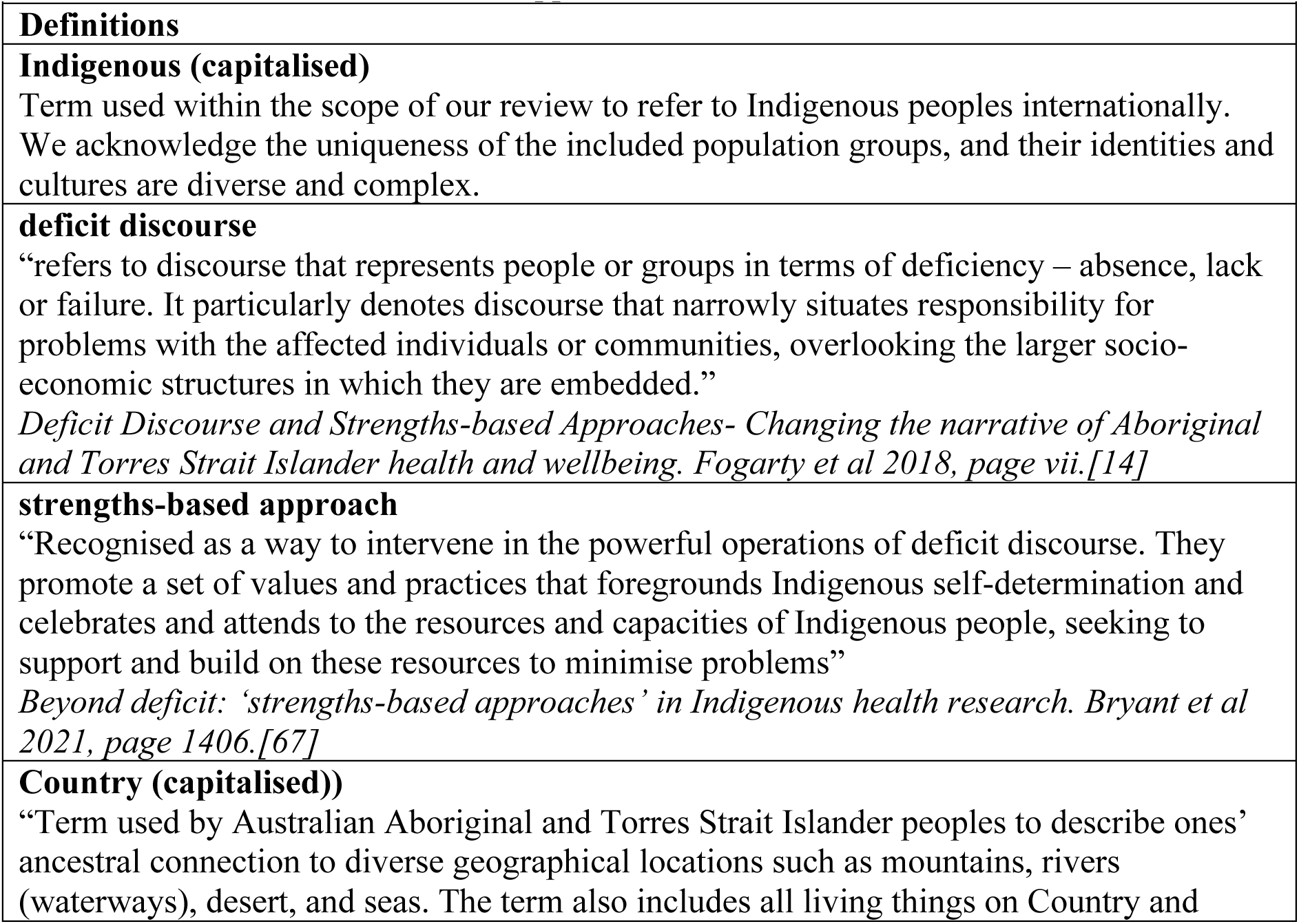
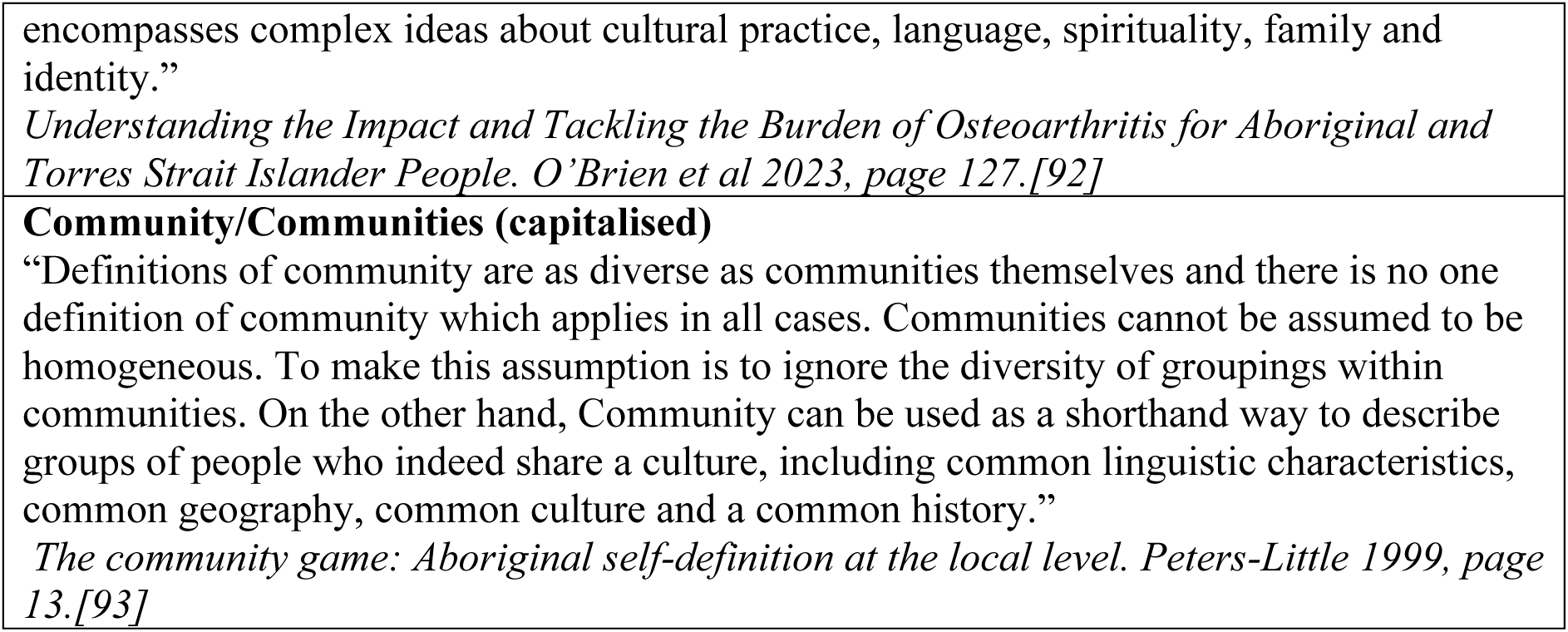

**Supplemental Table 2.**
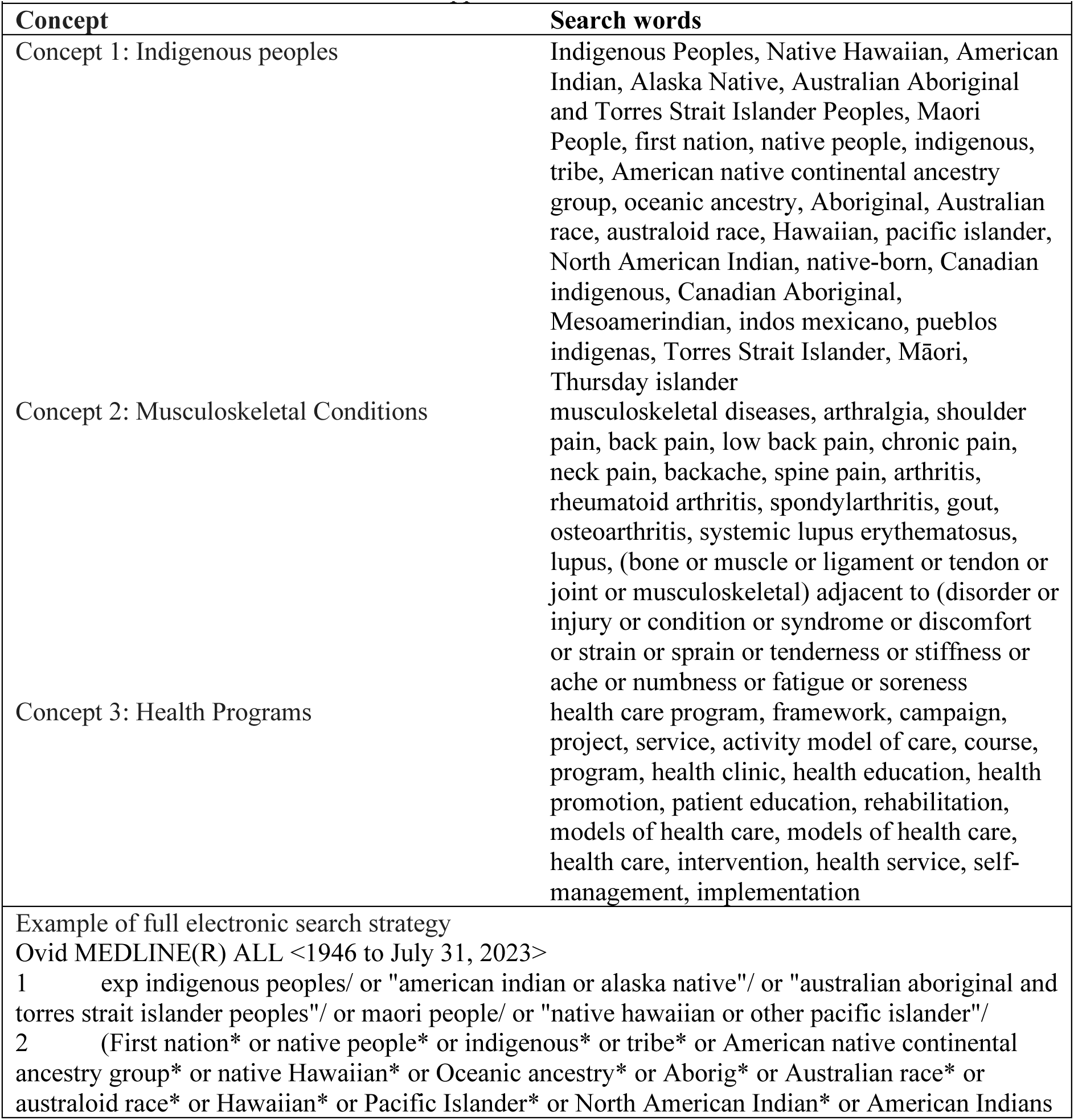
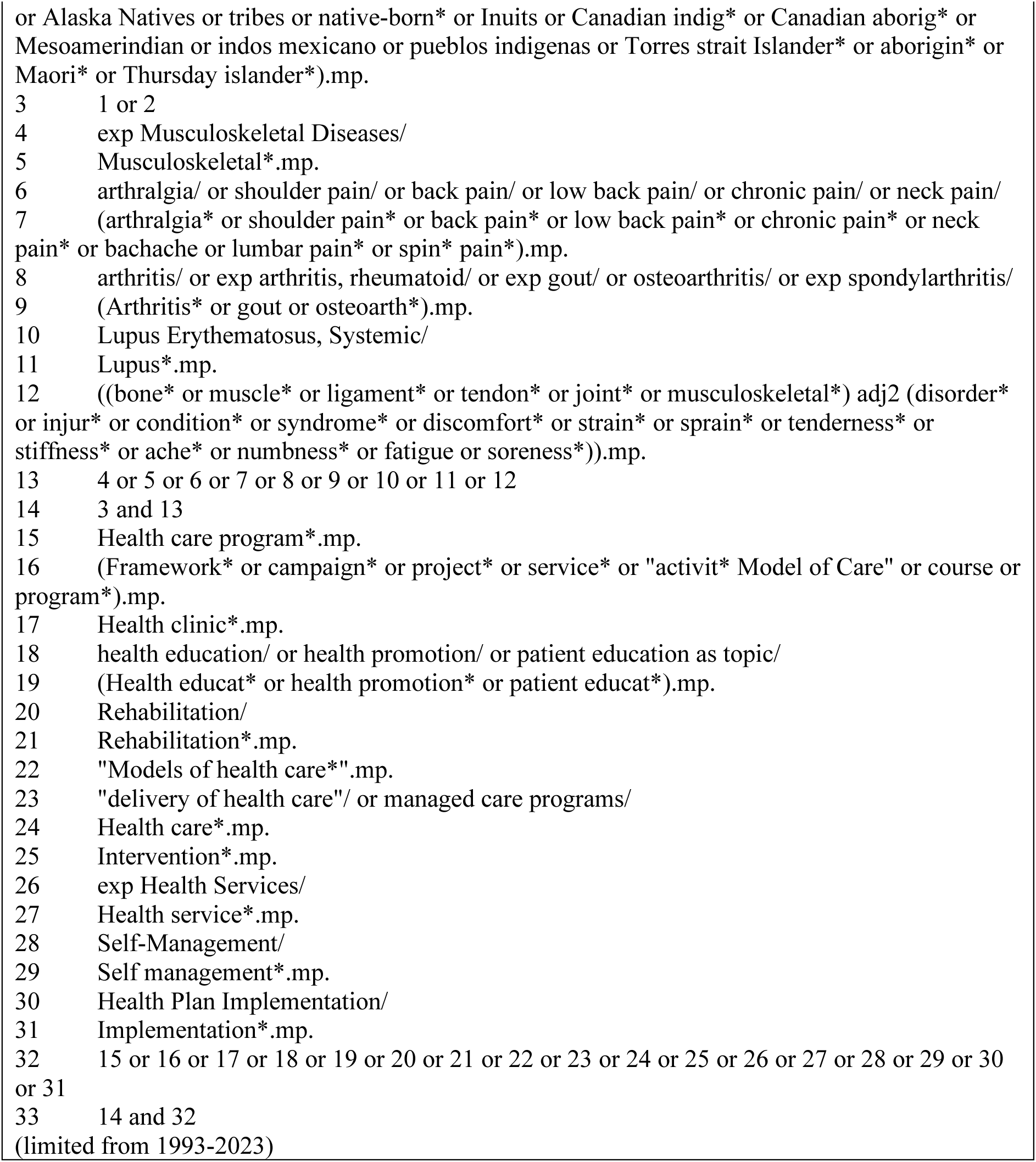

**Supplemental Table 3.**
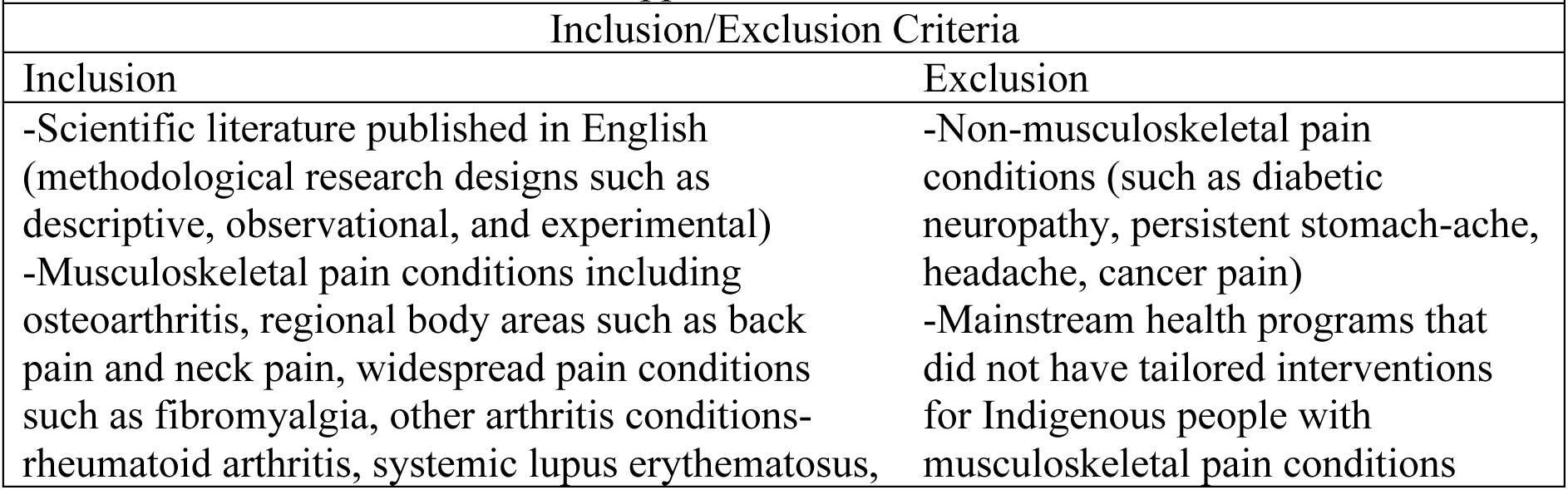
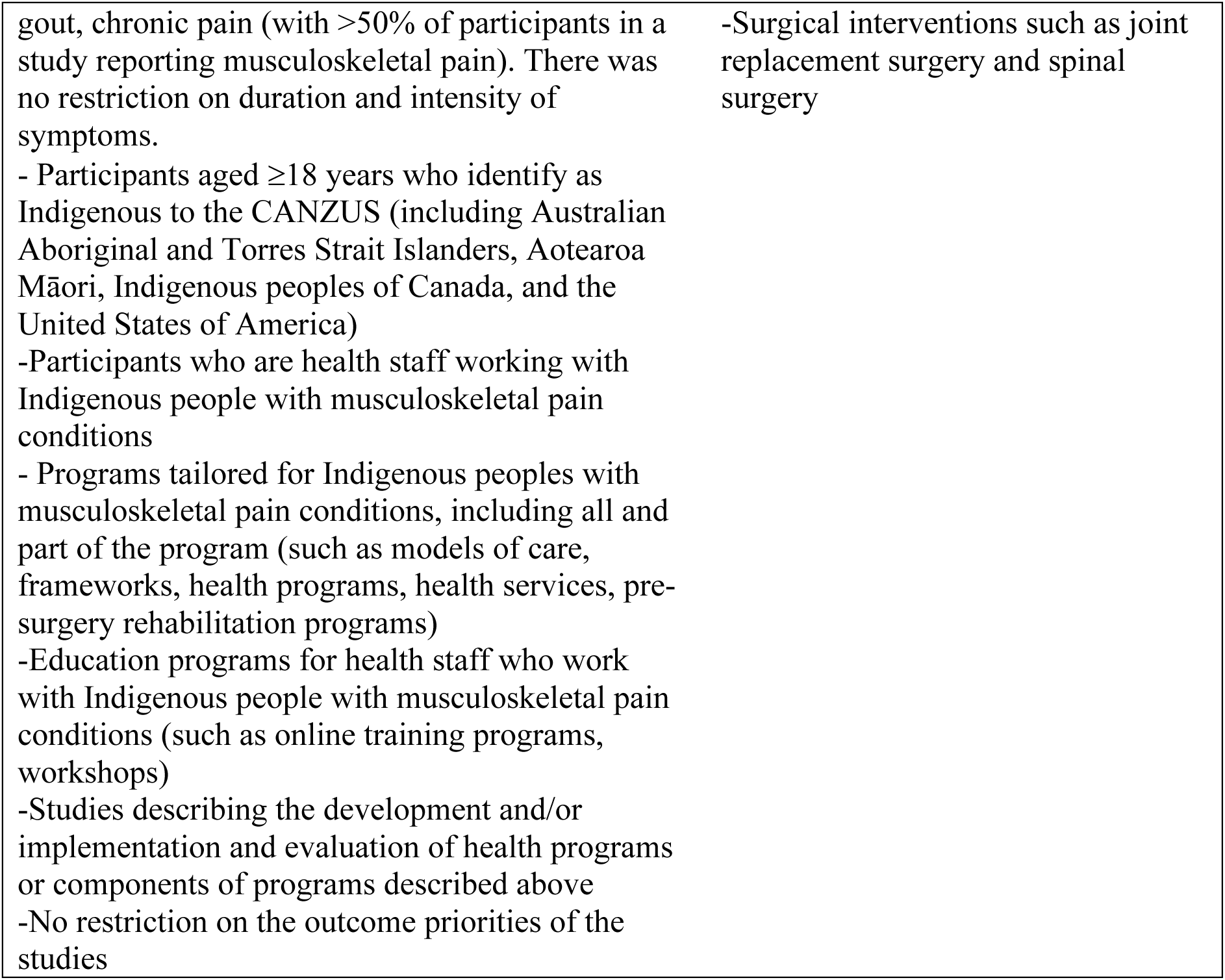

**Supplemental Table 4.**
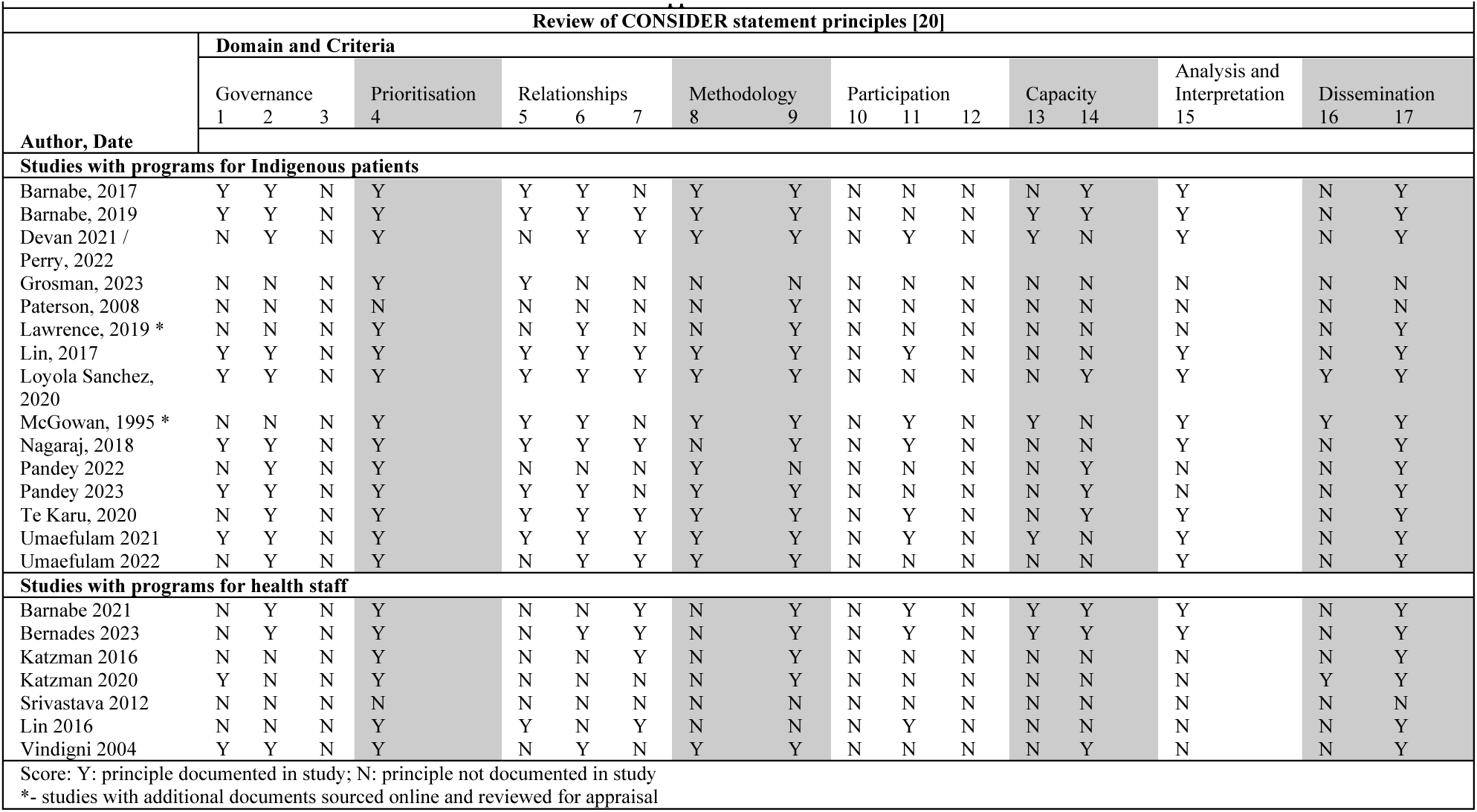

**Supplemental Table 5.**
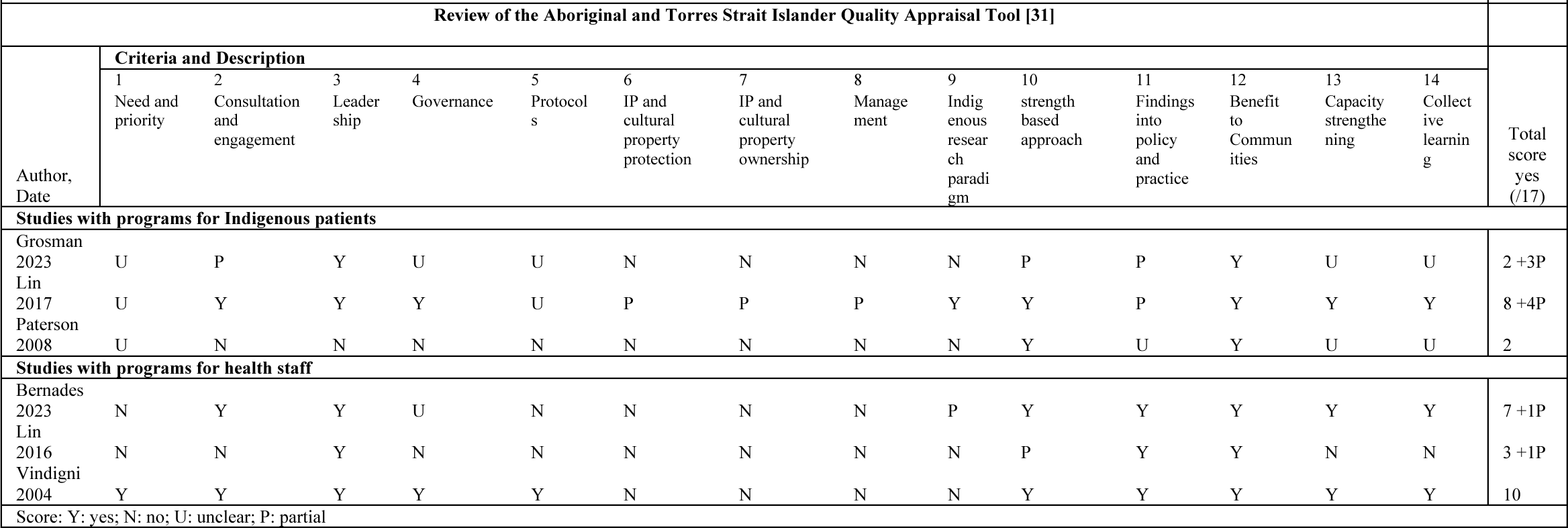

